# Predicting the risk of motor vehicle crash in the first year after cardioverter-defibrillator implantation

**DOI:** 10.1101/2025.03.05.25323435

**Authors:** John A Staples, Daniel Daly-Grafstein, Mayesha Khan, Shannon Erdelyi, Nathaniel M Hawkins, Herbert Chan, Christian Steinberg, Andrew D Krahn, Jeffrey R Brubacher

## Abstract

**Background:** Baseline health and driving data might allow clinicians to personalize medical driving restrictions after implantable cardioverter-defibrillator (ICD) implantation.

**Methods:** Using 22 years of population-based administrative data from British Columbia, Canada, we identified licensed drivers with a first ICD implantation between 1998 and 2018. After stratifying by ICD indication (primary vs secondary prevention of sudden cardiac death), we applied regression techniques to baseline health and driving data to estimate each driver’s 1- year crash risk. We assessed optimism-corrected discrimination and calibration of the final model using 200 bootstrapped samples.

**Results:** In the first year after implantation, there were 352 crashes among 3652 primary prevention ICD recipients and 270 crashes among 3408 secondary prevention ICD recipients. Crash prediction models exhibited poor discrimination (c-statistics 0.60 and 0.61, respectively) but good calibration (calibration slopes 1.14 and 1.07). The strongest predictors of crash among primary prevention ICD recipients were male sex, active vehicle insurance in the past year, and the number of crashes in the past year. The strongest predictors of crash among secondary prevention ICD recipients were male sex, no history of seizure, an active prescription for opioids, and active vehicle insurance in the past year.

**Conclusions:** Crash prediction models based on health and driving data had a limited ability to distinguish individuals who subsequently crashed from individuals who did not. Observed crash risks are likely to be strongly influenced by unobserved changes in road exposure (the hours or miles driven per week), limiting the application of these risk scores by clinicians and policymakers.

## Background

In the United States, Canada and Europe, a total of about 265,000 individuals per year receive an implantable cardioverter-defibrillator (ICD) to reduce the risk of sudden cardiac death (SCD).^1,2,3^ Arrhythmogenic syncope and unexpected shocks are believed to occur with increased frequency in the first few weeks after ICD implantation, transiently raising the risk of sudden cardiac incapacitation (SCI) while driving.^4–8^ Motor vehicle crashes caused by sudden cardiac incapacitation have resulted in serious injuries and even death.^9–12^ Clinical guidelines thus recommend temporary cessation of driving after ICD implantation (**eTable 1)**.^13–16^ Harms of medical driving restrictions include social isolation, loss of income and reduced quality of life, suggesting compulsory driving restrictions should only be used for patients with an unacceptably high anticipated risk of crash.^17–20^

Current post-implantation driving restrictions are based on the narrow clinical circumstances thought to influence the risk of SCI (e.g., Was there a recent cardiac incapacitation? Is there a high risk of another arrhythmia?). Other aspects of an individual’s medical and driving history are not considered, and no empirically derived risk estimation tool is available to guide management. This is in striking contrast to the personalized risk stratification routinely used by clinical cardiologists in the management of atrial fibrillation^21^, hypercholesterolemia^22^, acute myocardial infarction^23^, and cardiac risk prior to noncardiac surgery^24^.

Several prior studies have estimated average crash risks after ICD implantation. A population-based retrospective observational cohort study recently reported that 9373 drivers with recent ICD implantation exhibited a crash rate lower than matched controls (crude incidence rate, 8.5 vs 10.5 crashes per 100 person-years; adjusted hazard ratio, 0.71; 95%CI, 0.61-0.83).^25^ The Antiarrhythmics versus Implantable Defibrillators (AVID) trial found a self-reported an annual crash incidence rate of 3.4% among 627 drivers with ICDs (lower than the 4.9% reported among the U.S. general population).^26^ An uncontrolled single-centre case series of 171 drivers undergoing secondary prevention ICD implantation found that 6% reported crashing at a median follow-up of 38 months.^27^ A multicentre prospective cohort study of 241 ICD recipients in Ireland found that only 2.1% reported receiving a shock that resulted in a crash at a mean follow-up of 27 months.^28^ A survey of 213 ICD recipients in Italy found an annual reported crash rate of only 1.1%.^29^ Another survey of 2741 ICD recipients in Denmark reported only one shock-related crash at a median follow-up of 2.3 years. ^30^ However, no prior study has used empirical data to estimate individualized crash risks, leaving clinicians and guideline committees with limited data to inform personalized driving restriction recommendations after ICD implantation.

Seeking to address this gap, we developed and validated a risk estimation tool that uses baseline health and driving history to estimate individualized crash risk in the year following ICD implantation.

## Methods

### Baseline health and driving data

Our study was set in British Columbia (BC), Canada. Near the study midpoint, BC reported 613 traffic injuries and 9.9 traffic fatalities per billion vehicle-kilometres.^31^ We obtained province-wide individual-level ICD implantation data from Cardiac Services BC’s cardiac device registries and linked it to population-based administrative health data that included hospitalization records, physician fee-for-service claims, outpatient prescription drug fills, and vital statistics death data (**eMethods 1**).^32,33,34^ Baseline medical and driving history was established using a 1-year lookback interval from the ICD implantation date (**eFigure 1**).

Comorbidities were considered present when corresponding diagnostic codes were present in data for ≥1 hospitalization (including hospitalization for ICD implantation) or ≥2 physician visits in the 1-year lookback interval (**eTable 2**). Active prescription medication use at implantation was inferred using dispensation date and days supplied of prescriptions filled at any community pharmacy in BC. We linked health data with province-wide driving history and crash data (**eMethods 1**).^34,35^ Driving history included data on driver license type, years since full license, traffic contraventions (traffic law violation or ‘ticket’) in the prior year, and crashes in the prior year that were attended by police or that resulted in an insurance claim (**eTable 2**).

### Multiple imputation of missing baseline data

Data was missing for several key variables including ICD indication (primary vs secondary prevention of sudden cardiac death; missing for 25% of cohort), left ventricular ejection fraction (LVEF; missing for 71%), and New York Heart Association (NYHA) symptom classification (missing for 79%). Device type (dual-chamber or cardiac resynchronization vs single-chamber ICD) was missing for 81% of the cohort with unknown ICD indication but missingness was minimal when ICD indication was known (**eMethods 1**). We used baseline cardiac device registry and administrative health data to impute ICD indication, LVEF, NYHA class and device type for 40 imputation sets. Our team previously validated a similar strategy to impute ICD indication with 90% sensitivity and 89% specificity.^36^ Each imputation set was then independently used to define the cohorts and create the regression models as described below, with the descriptive statistics, final regression coefficients, and measures of validation reflecting results pooled across all imputation sets within each cohort.

### Creating the primary and secondary prevention ICD cohorts

Our study included individuals with a first ICD implantation occurring in BC between 1 January 1998 and 31 October 2018. We excluded patients with a record of a prior ICD and patients who did not have an active driver license on the ICD implantation date. After imputing ICD indication, we assigned individuals within each imputation set to either the ‘**primary prevention ICD cohort**’ or the ‘**secondary prevention ICD cohort**’.

### Outcome: All crashes

The outcome of interest was the involvement of the ICD recipient as a driver in ≥1 crash in the first year after implantation that was attended by police and/or resulted in an insurance claim (dichotomous variable; 1=any eligible crash; 0=no eligible crash). We simplified the analysis and interpretation of results by ignoring censoring and competing events (e.g., license expiry, license suspension, subsequent cardiac device procedure, death) because our previous work found these events affected only about 7% of individuals in the first 6 months after ICD implantation.^25^ Descriptive data focused on the first eligible crash for individuals with multiple crashes in the year after ICD implantation.

### Identifying potential predictors for the global regression model

We developed a list of potential predictors of crash based on literature review, clinical experience, and our prior familiarity with these data (**eTable 2**). We limited the number of variables in our global model so that there were about 10 events per variable. The final sample size allowed us to include up to 30 variables in each global model. We used the same global model variables for both cohorts.

### Analysis

We performed all analyses separately in the primary prevention and secondary prevention ICD cohorts. We described baseline health and driving characteristics, described the crashes that occurred in the year after ICD implantation, and then fit a logistic regression model that used baseline characteristics to predict the probability of being involved in a crash as a driver in the year following ICD implantation. Due to the large number of potential predictors and moderate number of crash events, we performed regularization and variable selection using least absolute shrinkage and selection operator (LASSO) logistic regression.^37,38,39^ The level of L1 regularization was chosen by minimizing the 10-fold cross-validation error.

We assessed crash prediction model discrimination using the area under the receiver operating characteristic curve (AUC or c-statistic). We assessed crash prediction model calibration with the calibration plot, calibration slope and calibration-in the-large. We then dichotomized 1-year crash risk at three potentially clinically actionable ‘higher risk’ thresholds of ≥10%, ≥15%, and ≥20% (based on the observed crash rate of 10.5 crashes per 100 person-years among matched controls from a related cohort study^25^), and calculated sensitivity, specificity, positive predictive value, and negative predictive value for each cutoff. We validated model performance using optimism-corrected measures, calculated by repeating the multiple imputation and variable selection process in 200 bootstrap resamples^39^.

We repeated all analyses after stratifying by sex and performed sensitivity analyses with alternate cohorts (e.g., excluding recipients with unknown ICD indication), alternate outcomes (e.g., only considering crashes that resulted in injury or fatality), and alternate follow-up intervals (e.g., 3, 6, 9 and 60 months).

### Ethics

The University of British Columbia Clinical Research Ethics Board approved the study and waived requirements for individual consent (H16-02043). Data were de-identified before release to investigators. Access to data provided by the Data Stewards is subject to approval but can be requested for research projects through the Data Stewards or their designated service providers (**eMethods 1**). All inferences, opinions, and conclusions drawn are those of the authors and do not reflect the opinions or policies of the Data Stewards. It was not feasible to involve patient or the public during the design, conduct, reporting, interpretation, or dissemination of the study.

## Results

### Primary prevention ICD cohort

There were 3652 eligible individuals in the primary prevention ICD cohort (**Figure 1**). Median age was 66 years [Q1=58, Q3=73] and only 18% were female; data on race were not available (**Table 1**). As expected, cohort members frequently had evidence of prior heart failure (92%), three quarters had a reported LVEF <35%, and almost half had NYHA Class III or IV heart failure symptoms. Many had active prescriptions for antihypertensives (88%), beta-blockers (76%), ACE inhibitors or ARBs (72%), and mineralocorticoid receptor antagonists (45%). A majority held an active vehicle insurance policy at some point in the past year (85%), and crashes in the year prior to ICD implantation were relatively common (12%).

**Figure 1.**
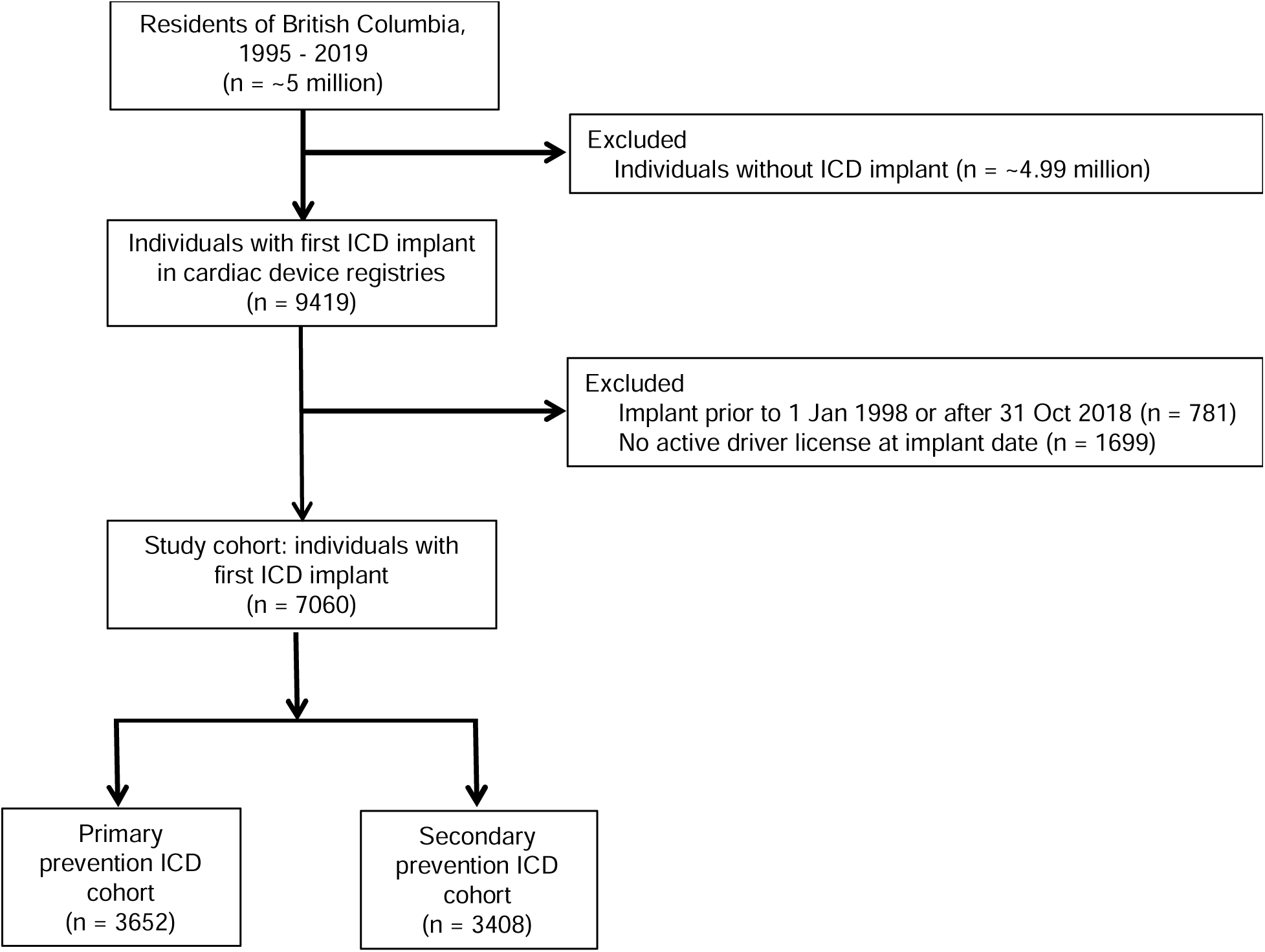
Study flow diagra. Using 40 multiple imputation sets, we assigned ICD indication (primary vs secondary prevention) for 25% of the sample who were missing this information. Each imputation set was used independently to define the primary prevention and secondary prevention cohorts, with results pooled across all imputation sets within each cohort.

**Table 1.**
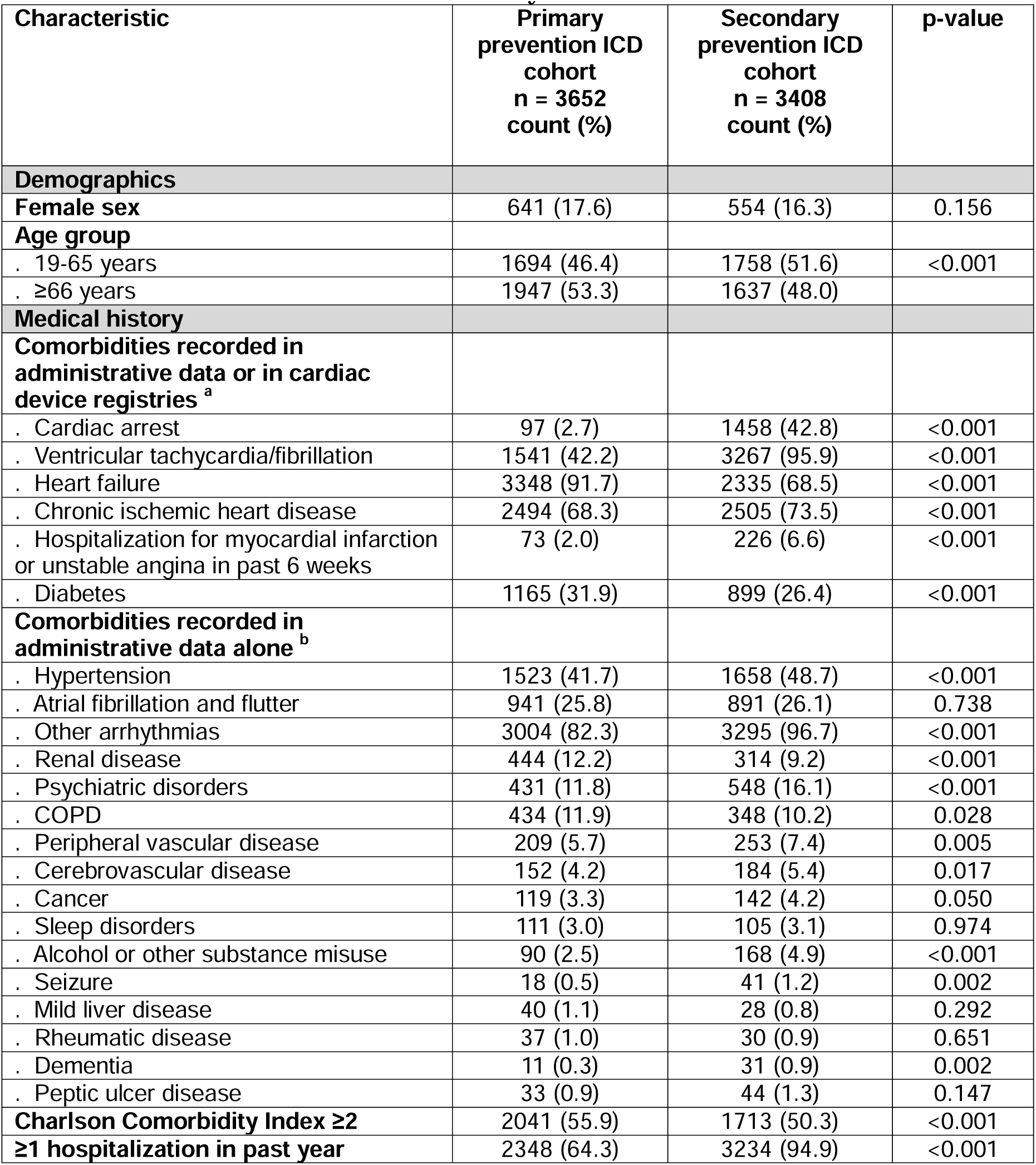
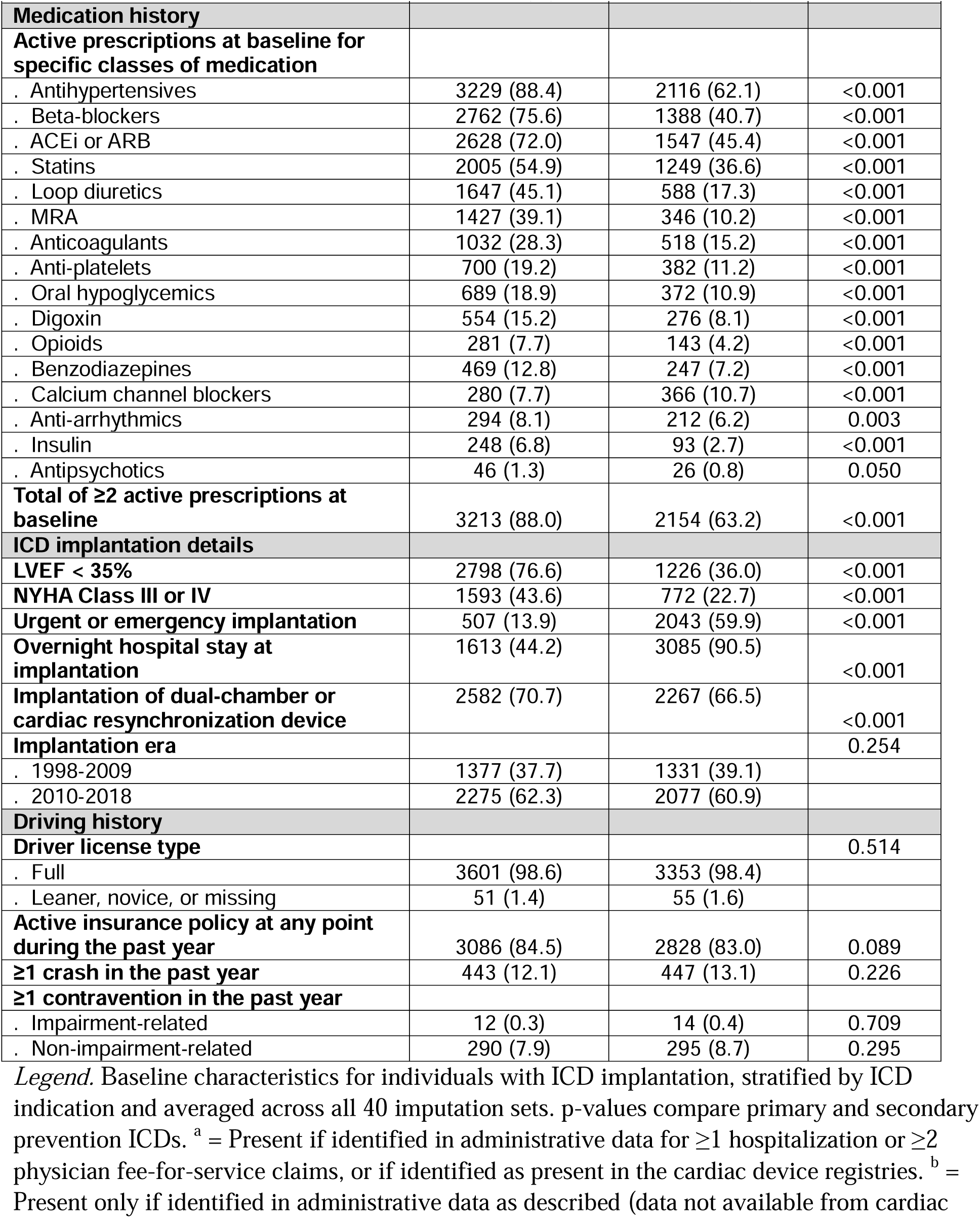
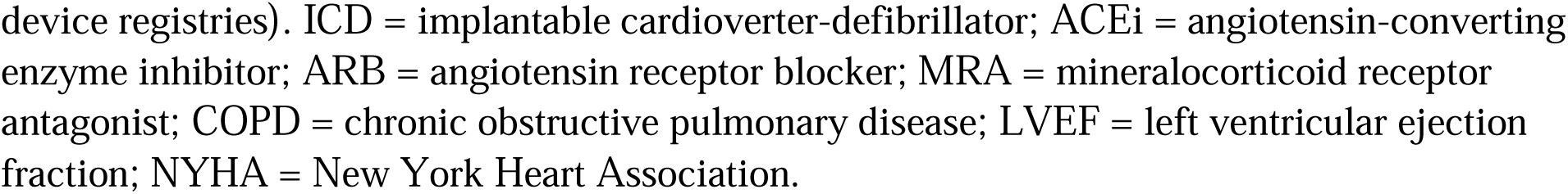
Baseline characteristics of the study cohorts. Baseline characteristics for individuals with ICD implantation, stratified by ICD indication and averaged across all 40 imputation sets. p-values compare primary and secondary prevention ICDs. ^a^ = Present if identified in administrative data for ≥1 hospitalization or ≥2 physician fee-for-service claims, or if identified as present in the cardiac device registries. ^b^ = Present only if identified in administrative data as described (data not available from cardiac device registries). ICD = implantable cardioverter-defibrillator; ACEi = angiotensin-converting enzyme inhibitor; ARB = angiotensin receptor blocker; MRA = mineralocorticoid receptor antagonist; COPD = chronic obstructive pulmonary disease; LVEF = left ventricular ejection fraction; NYHA = New York Heart Association.

A total of 352 (9.6%) of the 3652 drivers in the primary prevention ICD cohort crashed in the first year after ICD implantation. One in five crashes resulted in an injury or fatality (**eTable 3**). Among the 33 crashes attended by police, most involved ≥2 vehicles; alcohol, drugs, medications, and illness/fatigue were rarely listed by the attending officer as factors contributing to the crash (**eTable 4**).

The final primary prevention ICD cohort crash prediction model identified male sex, active vehicle insurance in the past year, and number of crashes in the past year as the strongest predictors of crash (**eTable 5**). The crash prediction model exhibited poor discrimination (c-statistic 0.60) but good calibration (calibration slope, 1.14; **Table 2**; **Figure 2**). Additional analyses found that model performance was similar in males and females (**eTable 7, eTable 8**) and that sensitivity analyses yielded results similar to those of the main analysis (**eTable 9**).

**Figure 2.**
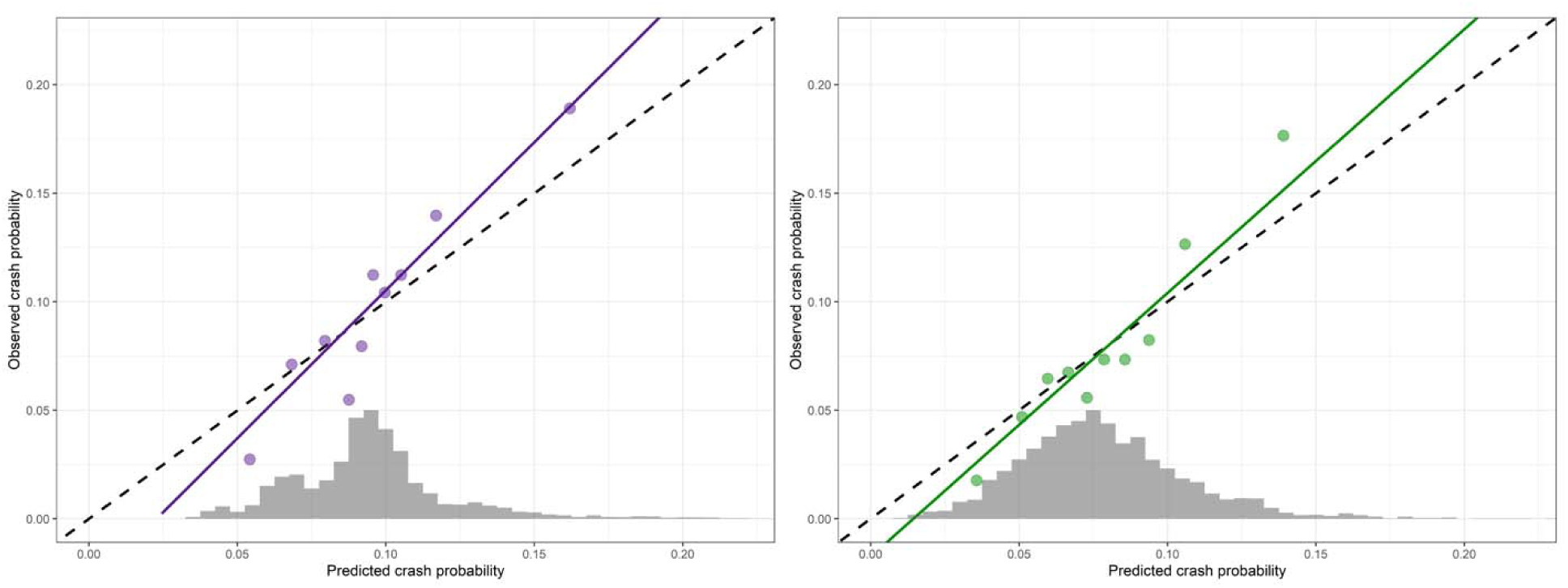
Distribution of predicted crash probabilities and calibration of crash prediction models. The figure on the left with purple depicts results from the primary prevention ICD cohort, while the figure on the right with green depicts results from the secondary prevention ICD cohort. The horizontal axis represents the predicted crash probabilities. The grey vertical bars are a histogram reflecting the distribution of predicted probabilities in each cohort, with bar height reflecting the proportion of each cohort in each group. Circular data points reflect the predicted and observed crash probabilities for predicted probability deciles of each cohort. The solid line reflects the optimism-corrected calibration slope for each prediction model, while the dashed line reflects perfect calibration. Main finding is that calibration appears to be excellent.

**Figure 3.**
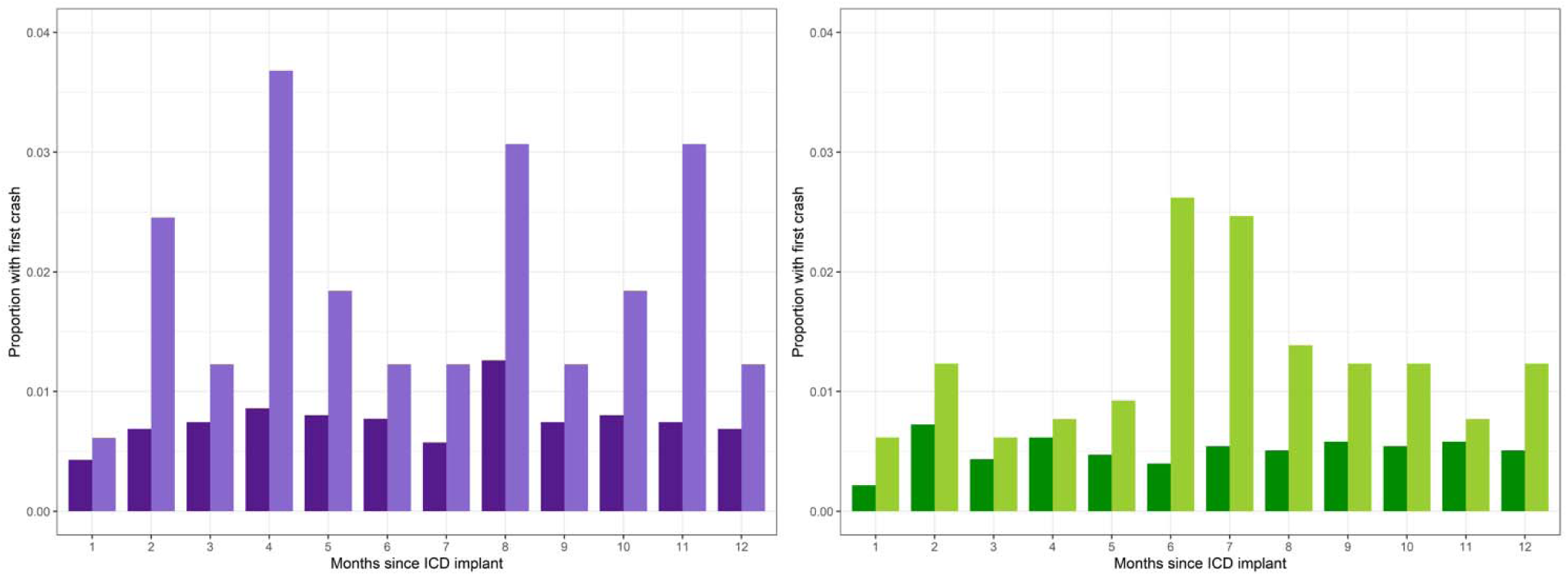
Crash probability after ICD implantation by month. The figure on the left with purple depicts results from the primary prevention ICD cohort, while the figure on the right with green depicts results from the secondary prevention ICD cohort. The horizontal axis represents the month after ICD implantation. The vertical axis represents the proportion of the cohort with a crash in the corresponding month. Darker vertical bars represent individuals with predicted crash probabilities of less than 15%, while lighter vertical bars represent individuals with predicted crash probabilities greater than 15%.

**Table 2.** Assessment of crash prediction model performance All values reflect the optimism-corrected average results pooled across 40 imputed data sets and were corrected for optimism using 200 bootstraps. Performance of dichotomized crash prediction models at clinically relevant thresholds (e.g., “≥10% cutoff” = ≥10% predicted risk of subsequent crash) illustrate the classic trade-off between sensitivity and specificity. AUC = area under the receiver operating curve.

| **Metric** | **Primary**<br>**prevention ICD**<br>**cohort**<br>**n = 3652** | **Secondary**<br>**prevention ICD**<br>**cohort**<br>**n = 3408** |
| --- | --- | --- |
| AUC | 0.60 | 0.61 |
| Calibration-in-the-large | 0.99 | 0.99 |
| Calibration slope | 1.14 | 1.07 |
| Sensitivity ($\geq 10\%$ cutoff) | 0.46 | 0.32 |
| Sensitivity ($\geq 15\%$ cutoff) | 0.08 | 0.05 |
| Sensitivity ($\geq 20\%$ cutoff) | 0.02 | 0.01 |
| Specificity ($\geq 10\%$ cutoff) | 0.66 | 0.82 |
| Specificity ($\geq 15\%$ cutoff) | 0.96 | 0.98 |
| Specificity ($\geq 20\%$ cutoff) | 0.99 | 1.00 |
| Positive predictive value ($\geq 10\%$ cutoff) | 0.13 | 0.14 |
| Positive predictive value ($\geq 15\%$ cutoff) | 0.20 | 0.24 |
| Positive predictive value ($\geq 20\%$ cutoff) | 0.23 | 0.28 |
| Negative predictive value ($\geq 10\%$ cutoff) | 0.92 | 0.93 |
| Negative predictive value ($\geq 15\%$ cutoff) | 0.91 | 0.92 |
| Negative predictive value ($\geq 20\%$ cutoff) | 0.90 | 0.92 |

### Secondary prevention ICD cohort

There were 3408 eligible individuals in the secondary prevention ICD cohort (**Figure 1**). Median age was 65 years [Q1=56, Q3=72] and only 16% were female (**Table 1**). Relative to the primary prevention ICD cohort, the secondary prevention ICD cohort was 2-fold more likely to have evidence of prior ventricular tachycardia or fibrillation, 16-fold more likely to have had a cardiac arrest in the past year, and 4-fold more likely to have required an ‘urgent/emergency’ rather than ‘elective’ ICD implantation. Almost everyone had a full driver license (rather than a learner or novice driver license) and held an active insurance policy at some point in the past year. In the year prior to ICD implantation, 13% had a serious crash and 9% had evidence of a traffic contravention.

A total of 270 (7.9%) of the 3408 drivers in the secondary prevention ICD cohort crashed in the first year after ICD implantation. One in four of these crashes resulted in an injury or fatality (**eTable 3**). Most of the 25 crashes attended by police involved ≥2 vehicles, and most of these crashes were not attributed to human factors (e.g., intoxication, medication, illness, speed; **eTable 4**).

The strongest predictors of crash in the secondary prevention ICD cohort were male sex, the number of impairment-related contraventions in the past year, having held active vehicle insurance in the past year, the absence of a history of seizure, and an active prescription for opioids at baseline (**eTable 6**). The secondary prevention ICD cohort crash prediction model exhibited poor discrimination (c-statistic, 0.61) but good calibration (calibration slope 1.07; **Table 2**; **Figure 2**). Model performance was similar in males and females (**eTable 7, eTable 8**) and results of sensitivity analyses were similar to results of the main analysis (**eTable 9**).

Based on these findings, we created a web-based risk estimation tool that could be used by clinicians to inform discussions about clinical risk (https://stapleslab.shinyapps.io/mvc-icd-risk-shiny-app/).^40^

## Discussion

We used 22 years of population-based health and driving data and a cohort of 7060 ICD recipients to develop a risk estimation tool that might inform personalized clinical decision-making about medical driving restrictions after ICD implantation. A total of 622 crashes occurred in the first year after ICD implantation, with 10% of primary prevention ICD recipients and 8% of secondary prevention ICD recipients experiencing a crash in the first year. Our crash prediction models showed excellent calibration, suggesting they might be useful to inform driving safety discussions between clinicians and patients (e.g., “In a group of patients with your history, X% had a crash in the year after ICD implantation”). However, despite use of extensive baseline demographic, health, and driving data, our crash prediction models exhibited limited ability to distinguish individuals who subsequently crashed from individuals who did not, suggesting limited clinical utility as a tool to identify individuals who would benefit from more stringent driving restrictions after ICD implantation.

Notably, baseline demographics and driving history were stronger determinants of subsequent crash risk than medical history. This reality is unsurprising but potentially underappreciated by clinicians and guideline committees, who might overestimate the importance of medical incapacitation as a cause of crash due to cognitive biases including base rate neglect and the availability heuristic (i.e., sudden cardiac incapacitation is frightening and easily brought to mind, perhaps leading to overestimation of its frequency).^41,42^ This also may present challenges for use of crash risk prediction tools in clinical practice. Clinicians are unlikely to have access to data on license class, crash history, and contravention history except through patient self-report. Moreover, clinicians may not feel it is their responsibility to assess risk factors obtained from a driving history, and might feel professionally and perhaps ethically conflicted if their advocacy for patients (who often wish to continue driving) conflicts with information from crash risk factors they perceive as being outside their professional expertise or scope of responsibility (e.g., crash history, contravention history).^43–47^

Symptoms of advanced heart failure (for primary prevention ICD recipients) and a history of seizures (for secondary prevention ICD recipients) were both counterintuitively associated with a *reduced* risk of subsequent crash, despite plausibly *increasing* the risk of sudden incapacitation. This is most credibly because individuals with worse health status drive less, and the annual likelihood of crash is a product of ‘crash risk per hour of driving’ and ‘hours of driving’. This point is particularly relevant to the current study because many individuals with the highest risk of sudden cardiac incapacitation (e.g., recurrent, uncontrollable ventricular tachycardia) will cease driving altogether, and will thus appear to have a very low annual crash risk.^48,49^ Advising these individuals to return to driving “because your risk is low” would be potentially catastrophic.

To our knowledge, no prior study has attempted to predict the risk of motor vehicle crash after ICD implantation using empirical data. Other strengths of this study include: use of large population-based cohorts to improve generalizability; exclusion of individuals who lacked an active driver license at ICD implantation; use of empirical (rather than self-reported) crash data, thus avoiding outcome misascertainment due to recall and self-reporting biases; use of data from the modern ICD era during which anti-tachycardia pacing and other innovations have markedly reduced the incidence of shocks; the use of modern methods for dealing with missing data; and the use of currently recommended procedures for the derivation and validation of predictive scores.

The major limitation of our study was the lack of a direct measure of road exposure. The fundamental problem is that a substantial proportion of drivers limit their road exposure in the weeks and months after ICD implantation, either because they were instructed to do so or because they feel less confident to drive in the face of new health problems. Understanding which patients ceased driving and for how long is nearly impossible because our data seldom includes sufficient clinical detail to know exactly which guideline-based restriction applies, because clinicians often neglect to advise patients about the applicable driving restriction^50^, because patient adherence to restrictions is limited^30^, and because some individuals cease driving because of health issues even when there is no requirement to do so. Our study had other limitations. We used state-of-the-art methods to deal with missing data, but data were commonly missing for ICD indication, LVEF, NYHA class and device type and such methods require unverifiable assumptions about being missing at random. We were unable to discern which crashes (if any) were caused by sudden cardiac incapacitation. We did not study commercial drivers. Individuals might not report a crash to the police or to the insurer, but this is likely rare, particularly for more serious crashes. Our risk score has not yet been validated on external data.

Medical driving restrictions after ICD implantation can significantly limit quality of life. Use of baseline health and driving data to predict post-implantation crash risk and personalize driving restrictions is a strategy that has the potential to strengthen clinical practice. Future studies exploring this strategy might benefit from collection of, or linkage to, road exposure data, driving behaviour data, and detailed arrhythmia detection data, both at baseline and during the outcome interval.

## Supporting information

TRIPOD checklist

## Data Availability

Access to data provided by the Data Stewards is subject to approval but can be requested for research projects through the Data Stewards or their designated service providers. All inferences, opinions, and conclusions drawn are those of the authors and do not reflect the opinions or policies of the Data Stewards. The following data sets were used in this study: Cardiac Services BC (CSBC), Consolidation file (includes demographics, registry, and census geodata), Hospital Separations, Medical Services Plan (MSP), PharmaNet, Vital Events and Statistics - Deaths, BC Traffic Accident System (TAS), Insurance Corporation of British Columbia (ICBC) files including the Driver Experience table, the Exam table, the Contraventions table, and Claims data. Further information regarding these data sets can be found in the PopData project webpage: https://my.popdata.bc.ca/project_listings/19-127/collection_approval_dates

