## Supplementary Appendix for "Predicting the risk of motor vehicle crash in the first year after cardioverter-defibrillator implantation"

John A Staples MD MPH

Daniel Daly-Grafstein PhD

Mayesha Khan MA

Shannon Erdelyi MSc

Nathaniel M Hawkins MD

Herbert Chan PhD

Christian Steinberg MD

Andrew D Krahn MD

Jeffrey R Brubacher MD

eTable 1: Driving restrictions after ICD implantation (private/non-commercial driving)

| **Procedure** | **USA** | **Canada/**  **British Columbia** | **Europe** |
| --- | --- | --- | --- |
| Primary prevention ICD implantation | No driving for at least 1 week (AHA HRS 2007 update). | 2003 guideline: No driving for 4 weeks. Resumption of driving if: Heart failure is NYHA class I, II, or III; regular clinic check with no malfunction; no impaired consciousness or disability due to therapy.  2023 guidelines: No driving for 1 week. | No driving for 4 weeks. |
| Secondary prevention ICD implantation,  initial sustained ventricular tachycardia WITHOUT impaired level of consciousness | No driving for 6 months. | 2003 guideline: No driving for at least 1 week; refrain from driving for 1 month from time of VT if LVEF≧30; refrain from driving for 3 months from time of VT if LVEF<30. Resumption of driving if heart failure is NYHA class I, II, or III.  2023 guidelines: No driving for 1 week after implant or appropriate ICD shock/therapy. | No driving for 3 months. |
| Secondary prevention ICD implantation, initial cardiac arrhythmia accompanied by impaired level of consciousness | No driving for 6 months. | 2003 guideline: No driving for 6 months after last episode of sustained symptomatic VT or syncope likely due to VT or cardiac arrest.  2023 guidelines: No driving for 3 months after last incapacitating event. | No driving for 3 months. |

*Legend.* ECG = electrocardiogram; AV = atrioventricular; VT = ventricular tachycardia; NYHA = New York Heart Association; LVEF = left ventricular ejection fraction; AHA HRS = American Heart Association Heart Rhythm Society. Adapted from prior publications.

eMethods 1: Data sources

A prior study used name, birthdate, provincial health insurance number and driver licensing number to probabilistically link cardiac devices registry data to population-based administrative health data and driving data. This had a linkage rate of >97%.37

| **Variables** | **Source of data** |
| --- | --- |
| **Health data** |  |
| ICD implantation data | Cardiac Services British Columbia (CSBC) cardiac device registries: the Cardiac Device Registry (CDR; 1998-2013) and HEARTis (2013-2019). |
| BC’s health insurance plan registration data | Consolidation File |
| Hospitalization data | Discharge Abstract Database (DAD) |
| Physician fee-for-service payment claim data | Medical Service Plan (MSP) |
| Outpatient prescription medication fill data | PharmaNet |
| Date and cause of death | Vital Statistics database |
| **Driving data** |  |
| Police-reported data for police-attended crashes | BC Traffic Accident System (TAS). Police in BC attend all fatal crashes, most serious injury crashes, and some crashes that resulted in property damage only. |
| Driver license data, contraventions data, motor vehicle insurance claims | Insurance Corporation of British Columbia (ICBC) files including the Driver Experience table, the Exam table, the Contraventions table, and Claims data. |

- **Cardiac Services BC**: Cardiac Services BC. Vancouver, BC: Provincial Health Services Authority, 2021. <http://www.cardiacbc.ca/>
  - Cardiac Devices registry (1995-2013); HEARTis registry (2013-2019)
- **Consolidation File**: British Columbia Ministry of Health [creator] (2020): Consolidation File (MSP Registration & Premium Billing). V2. Population Data BC [publisher]. Data Extract. MOH (2020). http://www.popdata.bc.ca/data
- **Discharge Abstract Database**: Canadian Institute for Health Information [creator] (2020): Discharge Abstract Database (Hospital Separations). V2. Population Data BC [publisher]. Data Extract. MOH (2020). http://www.popdata.bc.ca/data
- **Medical Services Plan**: British Columbia Ministry of Health [creator] (2020): Medical Services Plan (MSP) Payment Information File. Population Data BC [publisher]. Data Extract. MOH (2020). http://www.popdata.bc.ca/data
- **PharmaNet**: British Columbia Ministry of Health [creator] (2020): PharmaNet. V2. Population Data BC [publisher]. Data Extract. Data Stewardship Committee (2020). http://www.popdata.bc.ca/data
- **Vital Statistics**: British Columbia Ministry of Health [creator] (2020): Vital Events Deaths. V2. Population Data BC [publisher]. Data Extract. MOH (2020). http://www.popdata.bc.ca/data
- **Driver data** (Driver license, BC Traffic Accident System, ICBC Claims File): Insurance Corporation of British Columbia [creator] (2020): Driver Experience, Contraventions, and Exam tables and the Traffic Accident System. Insurance Corporation of British Columbia [publisher]. Data Extract. ICBC (2020).

All inferences, opinions and conclusions drawn in this manuscript are those of the authors and do not reflect the opinions or policies of the Data Stewards. Access to data provided by the Data Stewards is subject to approval, but can be requested for research projects through the Data Stewards or their designated service providers. Further information regarding these data sets can be found in the PopData project webpage: https://my.popdata.bc.ca/project_listings/19-127/collection_approval_dates

**Missing data**

- The current study used health and driving data that was linked probabilistically based on age and sex, so age and sex were never missing in the final linked data.
- If there was no crash recorded in TAS or ICBC Claims data, we inferred that no serious crash occurred. Thus, outcome was never missing.
- Many predictors were based on a qualifying event (e.g., prescription medication fill, hospitalization, clinic visit, traffic contravention). We coded these predictors as absent when the qualifying event was not present. Thus, many predictor variables were never missing.

However, data were incomplete, particularly for several important variables:

- ICD indication was ‘unknown’ for 1767 (25% of the full cohort) individuals with an active driver license at ICD implantation date.
- Left ventricular ejection fraction (LVEF) was missing for 5327 (75.5% of the full cohort) individuals.
- New York Heart Association (NYHA) symptom classification was missing for 5715 (80.9% of the full cohort) individuals.
- Device type was missing for 1433 (20.3% of the full cohort) individuals.

**Missingness stratified by ICD indication**

| **Variables** | **Indication known to be primary prevention**  **n=2804**  **count (%)** | **Indication known to be secondary prevention**  **n=2489**  **count (%)** | **Unknown indication for ICD**  **n=1767**  **count (%)** |
| --- | --- | --- | --- |
| Left ventricular ejection fraction (LVEF) | 2050 (73.1%) | 1605 (64.5%) | 1672 (94.6%) |
| New York Heart Association (NYHA) Class | 2082 (74.3%) | 1941 (78.0%) | 1692 (95.8%) |
| Device type | ≤5 | ≤5 | 1428 (80.8%) |

*Legend.* Data on LVEF and NYHA class were only available for the individuals included in the HEARTis era of the cardiac device registries (2013-2019). Data Stewards require that values ≤5 to be suppressed from publication for data privacy.

We performed multiple imputation for these missing data. A similar approach was used on these data to infer ICD indication for a prior study, and a validation study showed the algorithm had a ≥90% sensitivity and ≥89% specificity for secondary prevention ICDs.^[1]^

eFigure 1: Study schematic

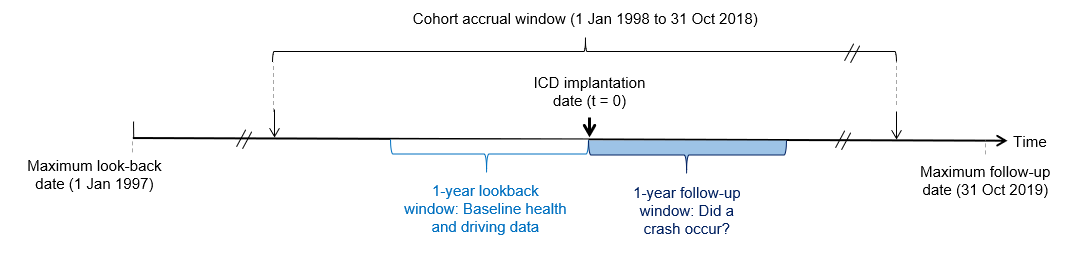

*Legend.* We used a 1-year lookback from implantation date to establish baseline health and driving history. The primary analysis assessed the risk of crash in the first year after ICD implantation.

eTable 2: Potential predictors for the global model

| **Predictor** | **Definition** |
| --- | --- |
| **Demographics** |  |
| Age | Driver age in years at implantation date (continuous). |
| Sex | Driver sex (male vs female*). |
| **Driving history** |  |
| License type | Type of driver license held at the time of ICD implantation (full vs 'novice or learner'*). |
| Active motor vehicle  insurance in the past year | Held active motor vehicle insurance at any point in the year before ICD implantation (yes vs no*). |
| Number of impairment-  related traffic contraventions  in the past year | Number of alcohol- or drug-impaired driving traffic violations in the year before ICD implantation (count). |
| Number of non-impairment-  related traffic contraventions  in the past year | Number of driving traffic violations that were not related to substance use in the year before ICD implantation (count). |
| Number of crashes in the  past year | Number of police-attended or insurance-claim crashes in the year before ICD implantation (yes vs no*). |
| **Medical history** |  |
| Number of overnight hospital  stays in the past year | Number of hospital discharges in the year before index date where DAD’s SEPDISP is ≥1 day after DAD’s ADDATE, including index hospitalization if present (yes vs no*). |
| History of cardiac arrest | Present (yes vs no*) if:   1) Diagnostic codes for “cardiac arrest” are present in ≥1 DAD or ≥2 MSP records in the past year; OR, 2) CSBC cardiac device registry variable indicating cardiac arrest.** |
| History of ventricular  tachycardia or ventricular  fibrillation (VT/VF) | Present (yes vs no*) if:   1) Diagnostic codes for “ventricular tachycardia and fibrillation” are present in ≥1 DAD or ≥2 MSP records in the past year; OR, 2) CSBC cardiac device registry variable indicating history of VT or VF.** |
| History of syncope | Present (yes vs no*) if:   1) Diagnostic codes for “syncope” are present in ≥1 DAD or ≥2 MSP records in the past year; OR, 2) CSBC cardiac device registry variable indicating syncope.** |
| Recent hospitalization for  myocardial infarction | Present (yes vs no*) if:   1) Discharge date of hospitalization with Most Responsible Diagnosis corresponding to “myocardial infarction” OR “unstable angina” ≤6 weeks prior to implantation date (for this variable, we did not look within MSP); OR, 2) CSBC cardiac device registry variable indicating “History of MI ≤6 weeks prior” (assoc_disease == “10. Myocardial Infarction <= 6 Weeks” in the CDR; no analogous variable in HEARTis).** |

**eTable 2: Potential predictors for the global model (continued)**

| **Predictor** | **Definition** |
| --- | --- |
| **Medical history (continued)** |  |
| History of heart failure | Present (yes vs no*) if:   1) Diagnostic codes for “congestive heart failure” are present in ≥1 DAD or ≥2 MSP records in the past year; OR, 2) CSBC cardiac device registry variable indicating heart failure.** |
| History of chronic ischemic  heart disease | Present (yes vs no*) if:   1) Diagnostic codes for either “myocardial infarction” OR “unstable angina” OR “chronic ischemic heart disease” are present in ≥1 DAD or ≥2 MSP records in the past year; OR, 2) CSBC cardiac device registry variable indicating chronic ischemic heart disease.** |
| History of diabetes | Present (yes vs no*) if:   1) Diagnostic codes for either “diabetes with complications” OR “diabetes without complications” are present in ≥1 DAD or ≥2 MSP records in the past year; OR, 2) CSBC cardiac device registry variable indicating diabetes.** |
| History of alcohol or  other substance misuse | Present if the diagnostic codes for “alcohol misuse” or “substance misuse” are present in ≥1 DAD or ≥2 MSP records in the past year (yes vs no*). |
| History of seizure disorder | Present if the diagnostic codes for “seizure disorder” are present in ≥1 DAD or ≥2 MSP records in the past year (yes vs no*). |
| History of obstructive sleep apnea and other sleep disorders | Present if the diagnostic codes for “obstructive sleep apnea and other sleep disorders” are present in ≥1 DAD or ≥2 MSP records in the past year (yes vs no*). |
| Left ventricular ejection fraction (LVEF) <35% | LVEF <35%, (vs LVEF ≥35%)*. Data on LVEF were only available from HEARTis (i.e.. the variable was not available in the CDR). LVEF data was already binned into an ordinal categorical (<20, 20-34, 35-50, 51-60, and >60) or binary (<35 vs ≥35) format in HEARTis, dictating the dichotomization threshold. Missing data were addressed using multiple imputation.** |
| NYHA class III or IV heart failure symptoms | Present if NYHA class III or IV (vs NYHA class I or II)*. Data on NYHA class were only available from HEARTis (i.e.. the variable was not available in the CDR). Missing data were addressed using multiple imputation.** |
| Baseline comorbidity burden  (CCI ≥2) | Charlson Comorbidity Index (CCI) ≥2, calculated using a diagnosis threshold of ≥1 DAD or ≥2 MSP claims in the past year for each component diagnosis (yes vs no*). |

**eTable 2: Potential predictors for the global model (continued)**

| **Predictor** | **Definition** |
| --- | --- |
| **Medical history (continued)** |  |
| Presence of selected  prescription medications | Present if medications from these classes were active at ICD implantation date, based on fill date and days supplied. Medications of interest included: benzodiazepines, opioids, and antipsychotics (each coded as a separate yes vs no* dichotomous indicator variable). |
| Number of active prescription  medications | Total number of unique active prescription medications at ICD implantation date (count). |
| **ICD Implantation details** |  |
| Urgent ICD implantation | 'Urgent/emergency’ implantation instead of ‘elective’ implantation*, as recorded in the cardiac device registries. |
| Overnight hospital stay at  implantation | For the ICD implantation visit, DAD’s SEPDISP date is one or more days after DAD’s ADDATE (yes vs no*). The index ICD implantation hospitalization was identified by comparing the implantation date and the ADDATE and SEPDISP dates for all of the individual’s hospitalizations (or for hospitalizations linked into a single episode of care). If no corresponding hospitalization was identified, we assumed there was no overnight hospital stay. |
| Type of ICD | Receipt of a dual-chamber ICD or cardiac resynchronization device (vs single-chamber ICD*), as recorded in the cardiac device registries. |
| **Other** |  |
| Calendar year of index date | Year of ICD implantation (continuous). |

*Legend.* * indicates the referent category used in regression analyses.

We were unable to include other comorbidities (e.g. atrial fibrillation or flutter; other arrhythmias; dementia; renal disease; liver disease (mild OR moderate or severe); cerebrovascular disease OR paraplegia and hemiplegia; cancer OR metastatic carcinoma; traumatic brain injury; psychiatric disorders) as predictors in the global model as doing so would cause the model to cross the recommended threshold of 10 events-per-variable. We excluded these variables because we believed they would not be strongly predictive of subsequent crash. Unless specifically described, all comorbidities were identified as present if the corresponding diagnostic codes were present in ≥1 DAD or ≥2 MSP records in the year prior to ICD implantation date.

DAD = Discharge Abstract Database. MSP = Medical Services Plan. LVEF = left ventricular ejection fraction. NYHA = New York Heart Association. CSBC = Cardiac Services British Columbia.

eTable 3: Characteristics of first crashes in the year after ICD implantation

| **Description** | **Crashes in the primary prevention ICD cohort,**  **count (%)**  **n = 352** | **Crashes in the secondary prevention ICD cohort,**  **count (%)**  **n = 270** | **p-value** |
| --- | --- | --- | --- |
| **Crash severity** |  |  |  |
| . Injury or fatality | 76 (21.6) | 72 (26.7) | 0.17 |
| . Property damage only | 276 (78.4) | 198 (73.3) |  |
| **Crash year** |  |  |  |
| . 1998-2005 | 41 (11.6) | 51 (18.9) | 0.04 |
| . 2006-2012 | 160 (45.5) | 107 (39.6) |  |
| . 2013-2018 | 151 (42.9) | 112 (41.5) |  |
| **Crash season** |  |  |  |
| . Winter (Dec - Feb) | 89 (25.3) | 66 (24.4) | 0.92 |
| . Spring (Mar - May) | 96 (27.3) | 79 (29.3) |  |
| . Summer (Jun - Aug) | 78 (22.2) | 55 (20.4) |  |
| . Fall (Sep - Nov) | 89 (25.3) | 70 (25.9) |  |
| **Crash day-of-week** |  |  |  |
| . Weekend (Fri - Sun) | 127 (36.1) | 120 (44.4) | 0.04 |
| . Weekday (Mon - Thurs) | 225 (63.9) | 150 (55.6) |  |
| **Crash time** |  |  |  |
| . Nighttime (21:00 - 6:00h) | 28 (8.0) | 21 (7.8) | 0.58 |
| . Daytime (6:00 - 18:00h) | 289 (82.1) | 215 (79.6) |  |
| . Evening (18:00 - 21:00h) | 35 (9.9) | 34 (12.6) |  |

*Legend*. For individuals with multiple crashes in the year after ICD implantation the earliest crash was taken as the unit of analysis.

eTable 4: Characteristics of police-attended crashes in the year after ICD implantation

| **Description** | **Police-attended crashes in the primary prevention ICD cohort,**  **count (%)**  **n = 33** | **Police-attended crashes in the secondary prevention ICD cohort,**  **count (%)**  **n = 25** | **p-value** |
| --- | --- | --- | --- |
| **Crash severity** |  |  | 0.02 |
| . Injury or fatality | 10 (30.3) | 16 (64.0) |  |
| . Property damage only | 23 (69.7) | 9 (36.0) |  |
| **Number of vehicles involved** |  |  | 1.00 |
| . Single vehicle | 7 (21.2) | 6 (24.0) |  |
| . Two or more vehicles | 26 (78.8) | 19 (76.0) |  |
| **Road location** |  |  | 1.00 |
| . City street | 24 (72.7) | 19 (76.0) |  |
| . Provincial highway or rural road | 9 (27.3) | 6 (24.0) |  |
| **Crash location** |  |  | 0.73 |
| . At intersection | 15 (45.5) | 9 (36.0) |  |
| . Between intersection | 8 (24.2) | 8 (32.0) |  |
| . Parking lot/other/unknown | 10 (30.3) | 8 (32.0) |  |
| **Human condition listed as contributing factor** |  |  | - |
| . Distracted/inattentive | 11 (33.3) | 11 (44.0) |  |
| . Illness/fatigue | ≤5 | ≤5 |  |
| . Alcohol | ≤5 | 0 (0.0) |  |
| . Drugs | 0 (0.0) | 0 (0.0) |  |
| . Medications | 0 (0.0) | 0 (0.0) |  |
| **Other contributing factors** |  |  | - |
| . Impaired by alcohol or drugs | ≤5 | ≤5 |  |
| . Excessive speed | ≤5 | ≤5 |  |
| **Speed zone** |  |  | - |
| . ≤ 50 km/h | 22 (66.7) | 14 (56.0) |  |
| . >50 km/h | 9 (27.3) | 8 (32.0) |  |
| . Other/unknown | ≤5 | ≤5 |  |
| **Weather** |  |  | - |
| . Clear/cloudy | 27 (81.8) | 17 (68.0) |  |
| . Rain/strong wind/snow/sleet/hail | 6 (18.2) | 7 (28.0) |  |
| . Fog/smoke/smog | 0 (0.0) | ≤5 |  |

*Legend*. Values ≤5 are suppressed from publication for data privacy according to the requirements of the Data Stewards. Chi-square tests were not run for groups with values ≤5. For individuals with multiple crashes in the year after ICD implantation the earliest crash was taken as the unit of analysis. When assessed as a proportion of all crashes, crashes resulting in injury or fatality were no more common among secondary prevention ICD recipient crashes than among primary prevention ICD recipient crashes (10 of 352 crashes vs 16 of 270 crashes, p=0.09).

eTable 5: Final prediction model effect estimates and variable selection stability metrics for the primary prevention ICD cohort

| **Predictors** | **Odds ratio (estimated 95%CI)** | **Imputation inclusion frequency** |
| --- | --- | --- |
| Intercept | 0.11 (0.06, 0.30) | 1.00 |
| Sex | 1.43 (1.12, 2.15) | 1.00 |
| Age | 0.99 (0.98, 1.00) | 1.00 |
| License type | 0.98 (0.39, 1.18) | 0.20 |
| Active insurance in past year | 1.57 (1.26, 2.44) | 1.00 |
| Number of impairment contraventions in past year | 1.04 (0.14, 2.42) | 0.15 |
| Number of non-impairment contraventions in past year | 1.15 (1.00, 1.42) | 1.00 |
| Number of crashes in past year | 1.50 (1.18, 1.86) | 1.00 |
| Number of overnight hospital stays in past year | 0.99 (0.83, 1.00) | 0.48 |
| History of cardiac arrest | 1.00 (0.63, 1.69) | 0.05 |
| History of ventricular tachycardia or ventricular fibrillation | 1.00 (0.93, 1.23) | 0.08 |
| Recent hospitalization for myocardial infarction | 1.03 (0.69, 2.18) | 0.30 |
| History of heart failure | 1.00 (0.70, 1.23) | 0.05 |
| History of chronic ischemic heart disease | 1.00 (0.95, 1.34) | 0.13 |
| History of diabetes | 1.00 (0.89, 1.22) | 0.03 |
| History of alcohol or other substance misuse | 0.97 (0.43, 1.15) | 0.30 |
| History of seizure | 1.00 (0.10, 2.09) | 0.00 |
| History of obstructive sleep apnea | 1.01 (0.77, 1.90) | 0.25 |
| Left ventricular ejection fraction <35% | 1.05 (1.00, 1.28) | 0.55 |
| NYHA class III or IV heart failure symptoms | 0.85 (0.70, 0.91) | 0.80 |
| CCI ≥2 | 1.00 (0.93, 1.25) | 0.00 |
| Presence of opioids at baseline | 1.00 (0.71, 1.22) | 0.00 |
| Presence of benzodiazepines at baseline | 1.00 (0.82, 1.22) | 0.00 |
| Presence of antipsychotics at baseline | 1.00 (0.35, 1.68) | 0.00 |
| Number of active prescriptions at baseline | 1.00 (0.99, 1.03) | 0.03 |
| Urgent/emergency ICD implantation | 1.00 (0.75, 1.16) | 0.00 |
| Overnight hospital stay at implantation | 1.00 (0.90, 1.30) | 0.00 |
| Dual-chamber ICD or cardiac resynchronization device | 0.99 (0.85, 1.17) | 0.15 |
| Year since start of study interval | 0.98 (0.95, 1.00) | 0.98 |

*Legend.* Displayed odds ratio values represent pooling results across all 40 imputation sets. The estimated 95% confidence intervals reflect the 2.5^th^ and 97.5^th^ quantile value of the estimated odds ratio among 200 bootstrap resamples. Imputation inclusion frequency denotes the proportion of imputation sets where the variable was included during the LASSO regression variable selection procedure. NYHA = New York Heart Association; CCI = Charlson Comorbidity Index.

eTable 6: Final prediction model effect estimates and variable selection stability metrics for the secondary prevention ICD cohort

| **Predictors** | **Odds ratio (estimated 95%CI)** | **Imputation inclusion frequency** |
| --- | --- | --- |
| Intercept | 0.12 (0.04, 0.32) | 1.00 |
| Sex | 1.78 (1.26, 2.81) | 1.00 |
| Age | 0.99 (0.98, 1.00) | 1.00 |
| License type | 1.00 (0.58, 1.96) | 0.08 |
| Active insurance in past year | 1.55 (1.17, 2.59) | 1.00 |
| Number of impairment contraventions in past year | 2.29 (0.88, 12.52) | 1.00 |
| Number of non-impairment contraventions in past year | 1.05 (0.89, 1.36) | 0.90 |
| Number of crashes in past year | 1.17 (1.00, 1.51) | 1.00 |
| Number of overnight hospital stays in past year | 1.15 (1.03, 1.32) | 1.00 |
| History of cardiac arrest | 1.00 (0.80, 1.15) | 0.23 |
| History of ventricular tachycardia or ventricular fibrillation | 0.89 (0.50, 1.24) | 0.78 |
| Recent hospitalization for myocardial infarction | 1.00 (0.74, 1.40) | 0.05 |
| History of heart failure | 1.00 (0.80, 1.21) | 0.18 |
| History of chronic ischemic heart disease | 0.98 (0.71, 1.09) | 0.50 |
| History of diabetes | 1.22 (1.00, 1.70) | 1.00 |
| History of alcohol or other substance misuse | 1.00 (0.57, 1.35) | 0.05 |
| History of seizure | 0.24 (0.09, 0.32) | 1.00 |
| History of obstructive sleep apnea | 1.00 (0.51, 1.48) | 0.05 |
| Left ventricular ejection fraction <35% | 1.01 (0.91, 1.15) | 0.50 |
| NYHA class III or IV heart failure symptoms | 0.95 (0.80, 1.04) | 0.68 |
| CCI ≥2 | 1.01 (0.86, 1.31) | 0.23 |
| Presence of opioids at baseline | 1.64 (1.01, 2.83) | 1.00 |
| Presence of benzodiazepines at baseline | 1.01 (0.77, 1.52) | 0.15 |
| Presence of antipsychotics at baseline | 0.85 (0.14, 1.30) | 0.48 |
| Number of active prescriptions at baseline | 0.96 (0.91, 0.99) | 1.00 |
| Urgent/emergency ICD implantation | 0.82 (0.60, 0.99) | 1.00 |
| Overnight hospital stay at implantation | 0.83 (0.54, 1.05) | 0.88 |
| Dual-chamber ICD or cardiac resynchronization device | 1.03 (0.92, 1.29) | 0.48 |
| Year since start of study interval | 0.99 (0.96, 1.00) | 0.95 |

*Legend.* See legend for eTable 5.

eTable 7: Assessment of crash prediction model performance, stratified by sex

a) Primary prevention ICD cohort

| **Metric** | **Males** | **Females** |
| --- | --- | --- |
| AUC | 0.58 | 0.58 |
| Calibration-in-the-large | 1.00 | 1.00 |
| Calibration slope | 1.14 | 3.56 |
| Sensitivity (≥10% cutoff) | 0.55 | - |
| Sensitivity (≥15% cutoff) | 0.09 | - |
| Sensitivity (≥20% cutoff) | 0.02 | - |
| Specificity (≥10% cutoff) | 0.56 | 0.99 |
| Specificity (≥15% cutoff) | 0.95 | 0.99 |
| Specificity (≥20% cutoff) | 0.99 | 1.00 |
| Positive predictive value (≥10% cutoff) | 0.13 | - |
| Positive predictive value (≥15% cutoff) | 0.19 | - |
| Positive predictive value (≥20% cutoff) | 0.21 | - |
| Negative predictive value (≥10% cutoff) | 0.91 | 0.93 |
| Negative predictive value (≥15% cutoff) | 0.90 | 0.94 |
| Negative predictive value (≥20% cutoff) | 0.90 | 0.94 |

b) Secondary prevention ICD cohort

| **Metric** | **Males** | **Females** |
| --- | --- | --- |
| AUC | 0.59 | 0.68 |
| Calibration-in-the-large | 0.99 | 0.96 |
| Calibration slope | 1.10 | 5.96 |
| Sensitivity (≥10% cutoff) | 0.34 | - |
| Sensitivity (≥15% cutoff) | 0.04 | - |
| Sensitivity (≥20% cutoff) | 0.01 | - |
| Specificity (≥10% cutoff) | 0.78 | 0.99 |
| Specificity (≥15% cutoff) | 0.98 | 0.99 |
| Specificity (≥20% cutoff) | 1.00 | 1.00 |
| Positive predictive value (≥10% cutoff) | 0.13 | - |
| Positive predictive value (≥15% cutoff) | 0.21 | - |
| Positive predictive value (≥20% cutoff) | 0.37 | - |
| Negative predictive value (≥10% cutoff) | 0.93 | 0.95 |
| Negative predictive value (≥15% cutoff) | 0.92 | 0.95 |
| Negative predictive value (≥20% cutoff) | 0.91 | 0.95 |

*Legend*. The table presents optimism corrected metrics. We were unable to perform some pre-specified sensitivity analyses for the female subgroup because no individuals had a predicted crash probability exceeding 10% (or 15%, or 20%), so sensitivities and positive predictive values for those cutoffs were undefined.

eTable 8: Prediction model effect estimates and variable selection stability metrics, stratified by sex

a) Primary prevention ICD cohort

|  | **Males** | | **Females** | |
| --- | --- | --- | --- | --- |
| **Predictors** | **Odds ratio**  **(estimated 95%CI)** | **Imputation inclusion frequency** | **Odds ratio**  **(estimated 95%CI)** | **Imputation inclusion frequency** |
| Intercept | 0.19 (0.10, 0.73) | 1.00 | 0.05 (0.00, 0.24) | 1.00 |
| Sex | 1.00 (1.00, 1.00) | 0.00 | 1.00 (1.00, 1.00) | 0.00 |
| Age | 0.99 (0.98, 1.00) | 1.00 | 1.00 (0.98, 1.02) | 0.00 |
| License type | 0.96 (0.29, 1.63) | 0.30 | 1.00 (0.32, 15.91) | 0.00 |
| Active insurance in past year | 1.37 (1.12, 2.51) | 1.00 | 1.30 (1.16, 43.50) | 0.58 |
| Number of impairment contraventions in past year | 1.03 (0.10, 3.29) | 0.15 | 1.00 (0.06, 1.00) | 0.00 |
| Number of non-impairment contraventions in past year | 1.15 (1.00, 1.48) | 1.00 | 1.00 (0.02, 1.62) | 0.00 |
| Number of crashes in past year | 1.48 (1.17, 1.83) | 1.00 | 1.05 (0.95, 3.48) | 0.33 |
| Number of overnight hospital stays in past year | 0.99 (0.81, 1.00) | 0.48 | 1.00 (0.76, 1.27) | 0.00 |
| History of cardiac arrest | 1.00 (0.55, 1.84) | 0.00 | 1.01 (0.07, 4.77) | 0.05 |
| History of ventricular tachycardia or ventricular fibrillation | 1.00 (0.85, 1.26) | 0.00 | 1.01 (0.72, 2.36) | 0.15 |
| Recent hospitalization for myocardial infarction | 1.04 (0.76, 2.19) | 0.38 | 1.00 (0.02, 0.94) | 0.00 |
| History of heart failure | 0.99 (0.59, 1.18) | 0.10 | 1.00 (0.72, 10.07) | 0.00 |
| History of chronic ischemic heart disease | 1.00 (0.89, 1.32) | 0.00 | 1.00 (0.72, 2.03) | 0.00 |
| History of diabetes | 1.00 (0.87, 1.29) | 0.05 | 1.00 (0.45, 2.34) | 0.00 |
| History of alcohol or other substance misuse | 1.00 (0.43, 1.29) | 0.08 | 1.00 (0.02, 0.92) | 0.00 |
| History of seizure | 1.00 (0.04, 1.74) | 0.00 | 1.00 (0.14, 1.00) | 0.00 |
| History of obstructive sleep apnea | 1.01 (0.72, 2.31) | 0.18 | 1.00 (0.02, 5.88) | 0.00 |
| Left ventricular ejection fraction <35% | 1.03 (1.01, 1.36) | 0.43 | 1.00 (0.68, 1.51) | 0.05 |
| NYHA class III or IV heart failure symptoms | 0.86 (0.68, 0.90) | 0.75 | 0.99 (0.73, 1.34) | 0.10 |
| CCI ≥2 | 1.00 (0.89, 1.39) | 0.03 | 1.00 (0.40, 1.33) | 0.00 |
| Presence of opioids at baseline | 1.00 (0.59, 1.25) | 0.00 | 1.00 (0.45, 3.00) | 0.00 |
| Presence of benzodiazepines at baseline | 1.00 (0.76, 1.31) | 0.00 | 1.00 (0.41, 1.54) | 0.00 |
| Presence of antipsychotics at baseline | 0.77 (0.05, 0.85) | 0.63 | 1.61 (0.12, 23.91) | 0.58 |
| Number of active prescriptions at baseline | 1.00 (0.99, 1.05) | 0.03 | 1.00 (0.91, 1.04) | 0.00 |
| Urgent/emergency ICD implantation | 1.00 (0.72, 1.18) | 0.00 | 1.00 (0.32, 1.50) | 0.00 |
| Overnight hospital stay at implantation | 1.00 (0.92, 1.33) | 0.00 | 1.00 (0.45, 1.44) | 0.03 |
| Dual-chamber ICD or cardiac resynchronization device | 1.00 (0.89, 1.24) | 0.03 | 0.91 (0.35, 1.03) | 0.38 |
| Year since start of study interval | 0.99 (0.95, 1.00) | 0.88 | 1.00 (0.88, 1.02) | 0.13 |

b) Secondary prevention ICD cohort

|  | **Males** | | **Females** | |
| --- | --- | --- | --- | --- |
| **Predictors** | **Odds ratio**  **(estimated 95%CI)** | **Imputation inclusion frequency** | **Odds ratio**  **(estimated 95%CI)** | **Imputation inclusion frequency** |
| Intercept | 0.17 (0.05, 0.59) | 1.00 | 0.07 (0.03, 5.41) | 1.00 |
| Sex | 1.00 (1.00, 1.00) | 0.00 | 1.00 (1.00, 1.00) | 0.00 |
| Age | 0.99 (0.98, 1.00) | 1.00 | 1.00 (0.96, 1.01) | 0.23 |
| License type | 1.00 (0.57, 4.92) | 0.05 | 1.01 (0.30, 6.02) | 0.13 |
| Active insurance in past year | 1.35 (1.09, 2.03) | 1.00 | 1.23 (1.16, 14.29) | 0.43 |
| Number of impairment contraventions in past year | 2.24 (0.95, 13.16) | 1.00 | 1.00 (1.00, 1.00) | 0.00 |
| Number of non-impairment contraventions in past year | 1.03 (0.89, 1.32) | 0.58 | 1.04 (0.40, 5.55) | 0.18 |
| Number of crashes in past year | 1.15 (0.96, 1.49) | 1.00 | 1.02 (0.72, 2.83) | 0.08 |
| Number of overnight hospital stays in past year | 1.16 (1.01, 1.36) | 1.00 | 1.00 (0.80, 1.68) | 0.00 |
| History of cardiac arrest | 1.00 (0.79, 1.14) | 0.13 | 1.00 (0.46, 1.78) | 0.00 |
| History of ventricular tachycardia or ventricular fibrillation | 0.99 (0.51, 1.40) | 0.23 | 0.89 (0.11, 1.28) | 0.28 |
| Recent hospitalization for myocardial infarction | 1.00 (0.64, 1.45) | 0.00 | 0.98 (0.22, 1.00) | 0.08 |
| History of heart failure | 1.01 (0.87, 1.26) | 0.13 | 0.94 (0.30, 1.14) | 0.35 |
| History of chronic ischemic heart disease | 1.00 (0.73, 1.18) | 0.10 | 0.95 (0.37, 1.78) | 0.20 |
| History of diabetes | 1.24 (1.01, 1.75) | 1.00 | 0.99 (0.19, 1.12) | 0.13 |
| History of alcohol or other substance misuse | 1.00 (0.67, 1.75) | 0.08 | 0.91 (0.10, 0.90) | 0.20 |
| History of seizure | 0.30 (0.10, 0.44) | 1.00 | 0.97 (0.03, 1.00) | 0.08 |
| History of obstructive sleep apnea | 1.00 (0.53, 1.37) | 0.03 | 1.03 (0.47, 11.58) | 0.10 |
| Left ventricular ejection fraction <35% | 1.03 (0.90, 1.18) | 0.43 | 0.91 (0.44, 1.09) | 0.13 |
| NYHA class III or IV heart failure symptoms | 0.97 (0.82, 1.05) | 0.50 | 0.89 (0.35, 0.86) | 0.20 |
| CCI ≥2 | 1.01 (0.89, 1.30) | 0.33 | 0.98 (0.34, 1.21) | 0.15 |
| Presence of opioids at baseline | 1.69 (1.01, 3.05) | 1.00 | 1.00 (0.33, 2.32) | 0.00 |
| Presence of benzodiazepines at baseline | 1.04 (0.86, 1.76) | 0.45 | 0.92 (0.11, 0.95) | 0.18 |
| Presence of antipsychotics at baseline | 0.93 (0.19, 1.82) | 0.30 | 0.99 (0.32, 1.00) | 0.03 |
| Number of active prescriptions at baseline | 0.96 (0.91, 0.99) | 1.00 | 1.00 (0.90, 1.02) | 0.08 |
| Urgent/emergency ICD implantation | 0.78 (0.59, 0.99) | 1.00 | 1.00 (0.74, 2.48) | 0.00 |
| Overnight hospital stay at implantation | 0.92 (0.59, 1.07) | 0.75 | 0.93 (0.16, 1.09) | 0.18 |
| Dual-chamber ICD or cardiac resynchronization device | 1.00 (0.87, 1.20) | 0.13 | 1.06 (0.92, 3.16) | 0.18 |
| Year since start of study interval | 0.99 (0.96, 1.00) | 0.85 | 1.00 (0.93, 1.04) | 0.00 |

*Legend*. NYHA = New York Heart Association; CCI = Charlson Comorbidity Index.

eTable 9: Results of sensitivity analyses

a) For primary prevention ICD cohort

|  | **AUC** | **Calibration-in-the-large** | **Calibration slope** |
| --- | --- | --- | --- |
| **Main analysis** | 0.63 | 1.00 | 1.39 |
| **Alternate cohort construction*** |  |  |  |
| . Exclude “ICD indication unknown”  (remaining n = 2804) | 0.60 | 0.99 | 1.11 |
| **Alternate outcome definitions**** |  |  |  |
| . Fatality or injury crashes only | 0.53 | 1.01 | 16.23 |
| . Single vehicle crashes only | 0.67 | 0.99 | 1.05 |
| **Alternate follow-up intervals** |  |  |  |
| . 3 months | 0.45 | 1.01 | 65.24 |
| . 6 months | 0.57 | 0.99 | 1.41 |
| . 9 months | 0.59 | 1.00 | 1.10 |
| . 12 months (main analysis) | 0.60 | 0.99 | 1.14 |
| . 5 years | 0.64 | 1.00 | 1.07 |

b) For secondary prevention ICD cohort

|  | **AUC** | **Calibration-in-the-large** | **Calibration slope** |
| --- | --- | --- | --- |
| **Main analysis** | 0.65 | 0.99 | 1.37 |
| **Alternate cohort construction*** |  |  |  |
| . Exclude “ICD indication unknown”  (remaining n = 2489) | 0.60 | 1.00 | 1.07 |
| **Alternate outcome definitions**** |  |  |  |
| . Fatality or injury crashes only | 0.63 | 0.98 | 1.61 |
| . Single vehicle crashes only | 0.58 | 0.98 | 2.91 |
| **Alternate follow-up intervals** |  |  |  |
| . 3 months | 0.58 | 1.01 | 2.02 |
| . 6 months | 0.59 | 0.99 | 1.44 |
| . 9 months | 0.61 | 0.99 | 1.10 |
| . 12 months (main analysis) | 0.61 | 0.99 | 1.07 |
| . 5 years | 0.63 | 1.00 | 1.04 |

*Legend*. * Subgroup analysis that only included ICD with established indication (i.e., it excluded ICDs with imputed indication only). ** Medical incapacitation might be more likely to result in a single-vehicle crash or casualty crashes (e.g., made no effort to brake or turn, drove off roadway, etc.).
